# Burst vs.tonic SCS for early zoster-associated pain: A retrospective cohort study

**DOI:** 10.1101/2025.08.01.25332733

**Authors:** Rixin Wen, Tongyu Liu, Kunming Peng, Meng Jia, Chenxing Li

## Abstract

**Objective:** Inadequate pain control during early zoster-associated pain (ZAP) can lead to postherpetic neuralgia (PHN), and traditional treatments have limitations. This retrospective analysis compares the analgesic effects of burst (BurstSCS) versus traditional tonic (TonicSCS) spinal cord stimulation in early ZAP patients, using retrospective data. It also evaluates differences in psychological state and sleep quality. The findings offer insights into the multidimensional effects of these SCS modalities for future research.

**Methods:** Retrospective analysis included 40 consecutive early ZAP patients undergoing SCS trial (From March 1, 2023 to March 1, 2025). Groups: BurstSCS (n=20) vs. TonicSCS (n=20) based on documented treatment selection. Outcomes assessed at baseline, 14, and 30 days: visual analog scale (VAS), Pittsburgh Sleep Quality Index (PSQI), Pain Vigilance and Awareness Questionnaire (PVAQ). Intergroup comparisons used independent t-tests (significant at P<0.05).

**Results:** At 14 days, BurstSCS achieved greater pain reduction (VAS: 1.95 ± 0.76 vs 2.65 ± 0.75; *P*=0.006) and better secondary outcomes:PSQI: 7.00 ± 2.08 vs 8.90 ± 1.89,PVAQ: 15.55 ± 2.80 vs 22.20±2.42,(all *P*<0.05). At 30 days, VAS was comparable (1.40±1.23 vs 1.80±0.95) but BurstSCS maintained lower PSQI (4.90±1.33 vs 5.85±1.60) and PVAQ (9.00±1.56 vs 18.50±2.16) (*P*<0.05).

**Conclusions:** BurstSCS provided superior early pain reduction (>70% VAS decrease at 14 days) and sustained improvements in sleep quality and pain vigilance in early ZAP patients. These findings support its potential for multidimensional symptom management, warranting validation through prospective trials. **KEYWORDS:**Spinal cord stimulation; Neuralgia, Postherpetic; Retrospective studies; Pain management.

## INTRODUCTION

Herpes zoster (HZ) affects approximately 1 million Americans annually, with global incidence rising due to aging populations and immunosuppressant use ^[1]^. Over 95% of adults harbor latent varicella-zoster virus (VZV)^[2]^; reactivation causes dorsal root ganglion inflammation^[3]^, leading to herpetic eruptions and zoster-associated pain (ZAP) ^[4,5]^.ZAP—encompassing acute pain and postherpetic neuralgia (PHN)—severely impairs quality of life ^[6,7]^. While pharmacotherapy is limited by side-effects (34.2% discontinuation rate ^[8]^), minimally invasive SCS mitigates central sensitization^[9]^ and reduces PHN incidence by 48% ^[10]^. Traditional TonicSCS shows suboptimal efficacy (47.3% responders ^[11]^) due to paresthesia discomfort ^[12]^ and incomplete coverage^[13]^.Burst SCS (BurstSCS) delivers clustered pulses with minimal paresthesia, potentially enhancing pain-emotion modulation ^[14-17]^. Despite promising case reports in acute ZAP ^[10,18]^, comparative effectiveness data against TonicSCS remain scarce. This retrospective analysis compared BurstSCS versus TonicSCS for multidimensional outcomes in early ZAP patients.

## METHODS

Retrospective comparative analysis of early ZAP patients receiving short-term SCS at Shenzhen Guangming District People’s Hospital (March 1, 2023 to March 1, 2025).This retrospective cohort study was conducted under ethical oversight (Approval No. LL-KT-2025077) with individual written consent.All data were securely stored and kept conffdential throughout the entire research process.

The patient screening process is shown in Fig.1. Of 62 consecutive early ZAP patients screened between March 2023 and March 2025, 22 were excluded (8 did not meet inclusion criteria, 14 due to patient refusal), leaving 40 patients included for analysis.

**Fig. 1:**
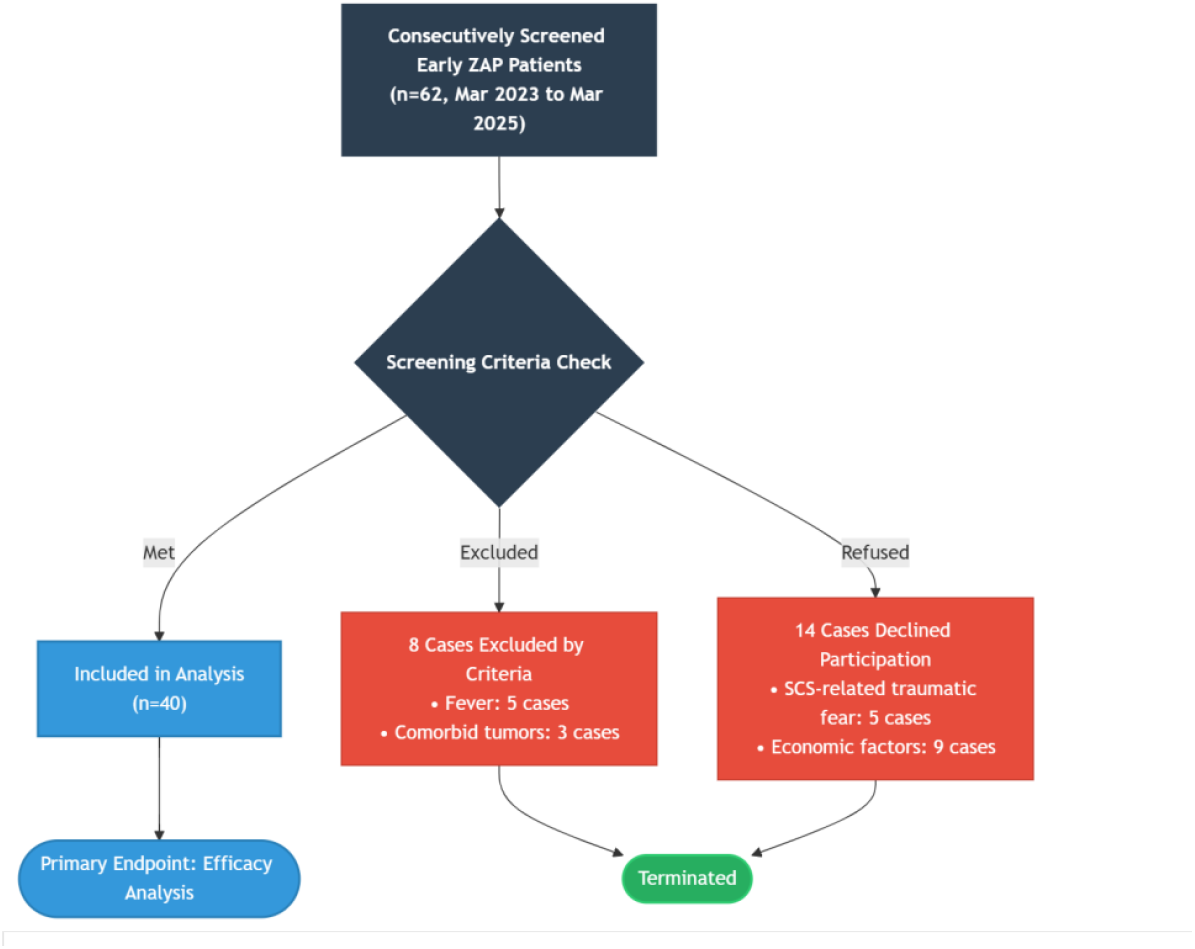
Patient enrollment flowchart

Patients were included if they had a confirmed diagnosis of HZ, defined by a unilateral dermatomal rash with neuropathic pain, with acute ZAP of ≤3 months’ duration and a baseline VAS score ≥6. Exclusion criteria were contraindications to SCS or sedation, coagulopathy (INR >1.5 or platelet count <100×10^9^/L), active infection or fever (>38°C), and psychiatric or cognitive disorders. Patients were assigned to either the BurstSCS group (n=20), with documented selection of burst stimulation, or the TonicSCS group (n=20), with documented selection of tonic stimulation. All patients received standard pharmacotherapy, including valacyclovir 500 mg three times daily for 7 days, pregabalin 75–150 mg twice daily for 14 days, and mecobalamin 0.5 mg three times daily for 30 days. All underwent SCS with percutaneous implantation of 8-contact electrodes, with the electrode tip placed at the affected dermatome level and removed on postoperative day 14. The primary outcome was VAS, and secondary outcomes included the PSQI and the PVAQ, assessed at baseline, day 14, and day 30. Continuous variables were expressed as mean ± standard deviation and analyzed using independent t-tests, while categorical variables were analyzed with χ^2^ tests. Group × time interactions were assessed using two-way analysis of variance (ANOVA), and effect sizes were calculated using Cohen’s d with 95% confidence intervals. A P-value <0.05 was considered statistically significant, and all analyses were performed using SPSS version 25.0.

## RESULTS

### Baseline Characteristics

The groups were foundto be similar in terms of age,sex,Disease duration and Affected segments (Table 1).

**Table 1.**
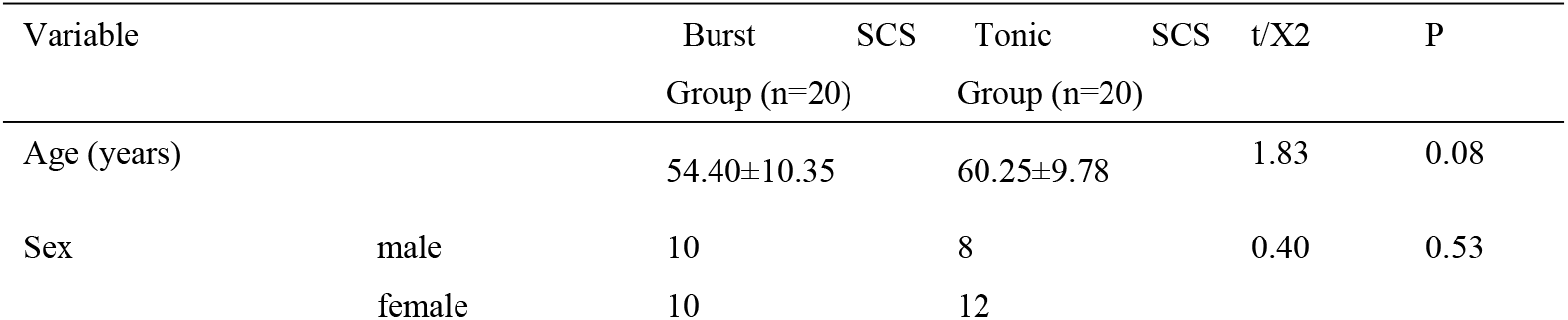
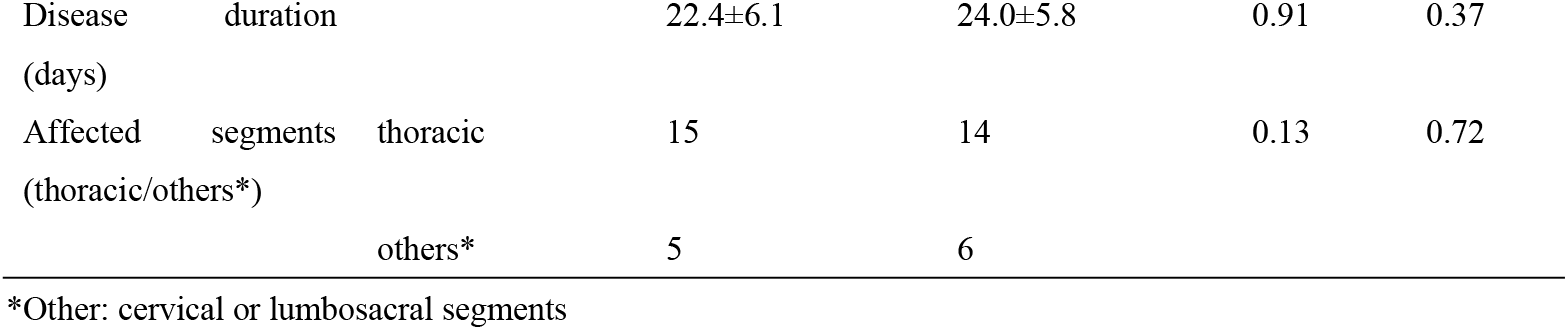
Baseline Characteristics Extracted from Medical Records

### Primary Outcome: VAS

BurstSCS demonstrated superior early analgesia (Table 2, Fig.2):

**Fig 2.**
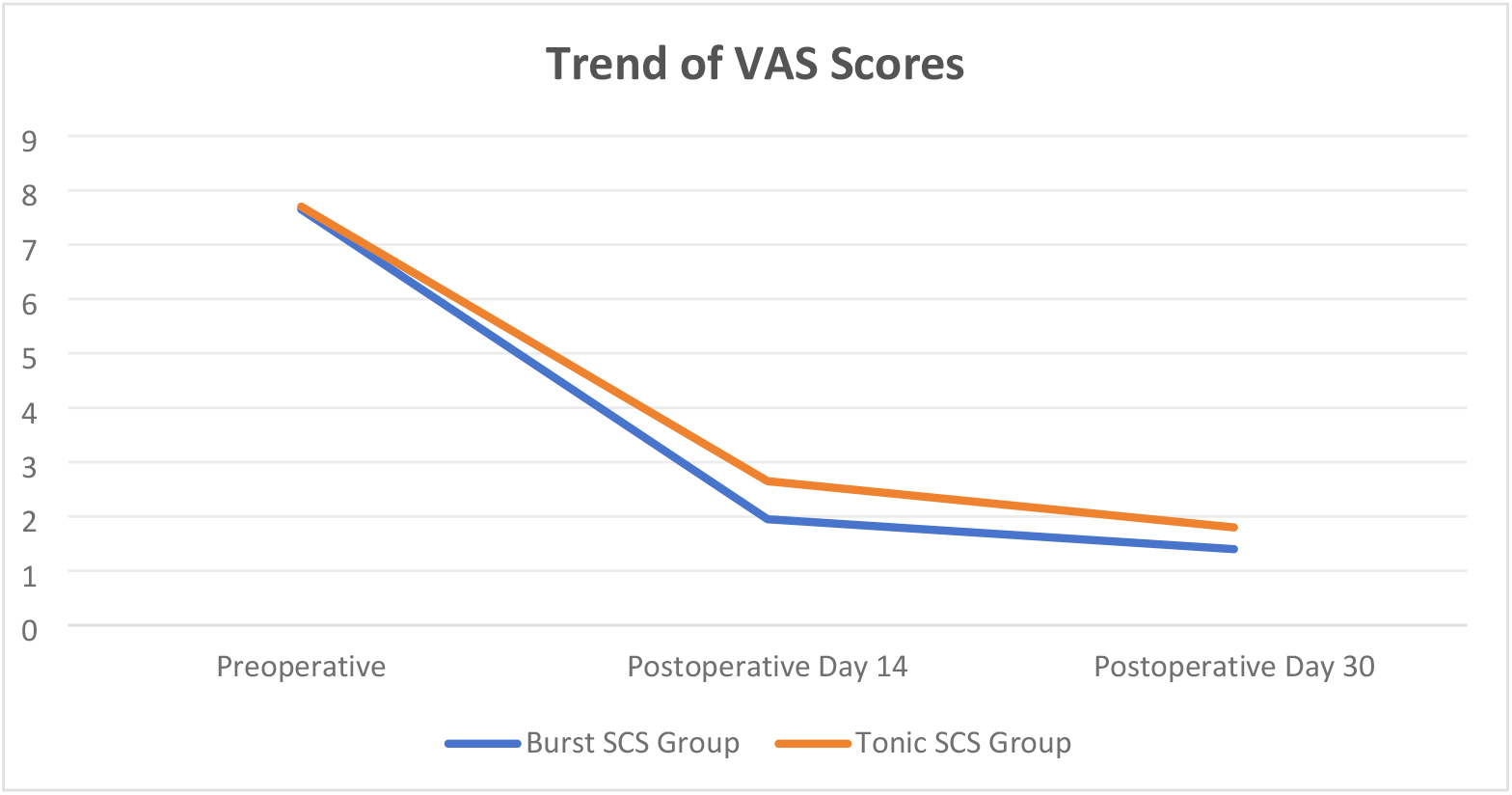
Documented VAS Score Trends

**Table 2.**
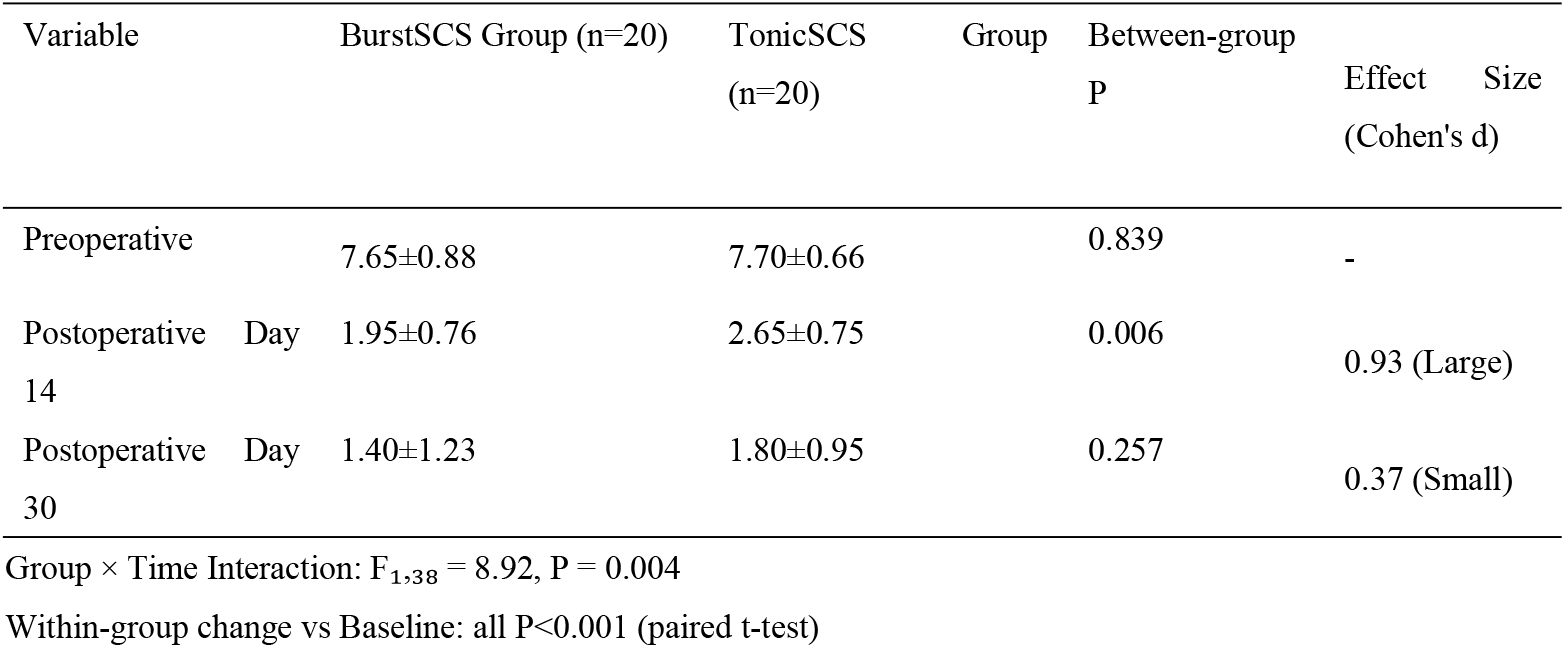
Documented VAS Score Comparisons

- Day 14 : 1.95±0.76 vs 2.65±0.75 (P=0.006), effect size d=0.93
- Day 30 : Comparable scores (1.40±1.23 vs 1.80±0.95, P=0.257)

Group×time interaction: P=0.004

### Secondary Outcomes

#### 1. PSQI

BurstSCS showed sustained improvement (Table 3, Fig.3):

**Fig 3.**
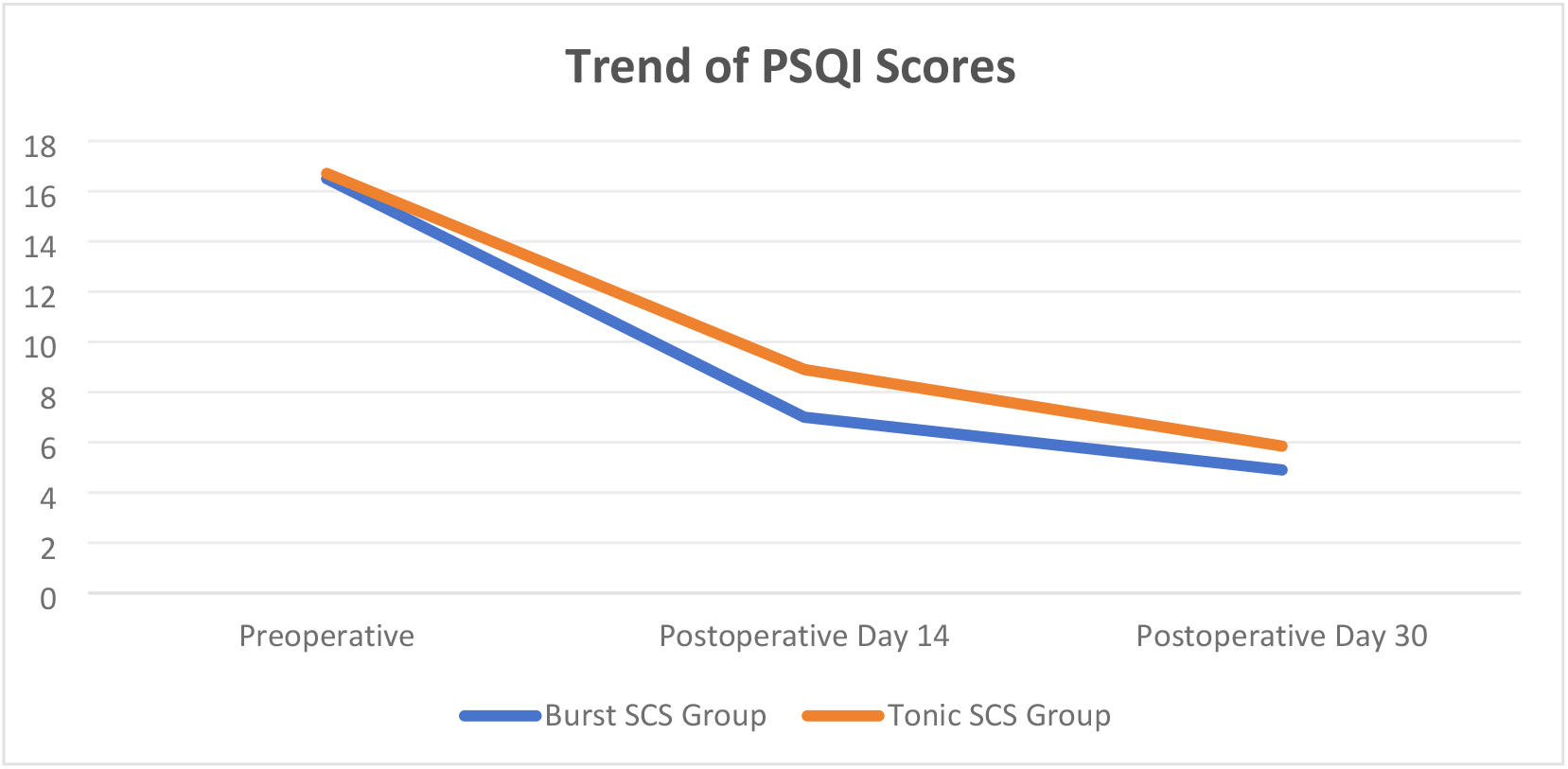
Documented PSQI Score Trends

**Table 3.**
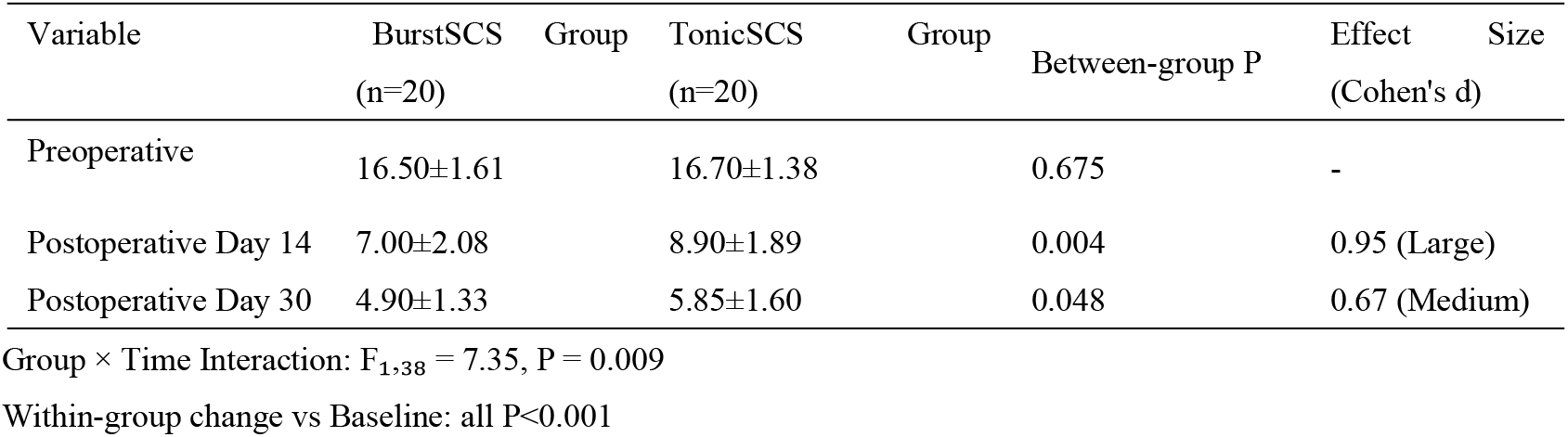
Documented PSQI Score Comparisons

- Day 14 : 7.00±2.08 vs 8.90±1.89 (P=0.004)
- Day 30 : 4.90±1.33 vs 5.85±1.60 (P=0.048)

Normative PSQI≤5 achieved by BurstSCS at Day 30

#### 2. PVAQ

Profound between-group differences favoring BurstSCS (Table 4, Fig.4):

**Fig 4.**
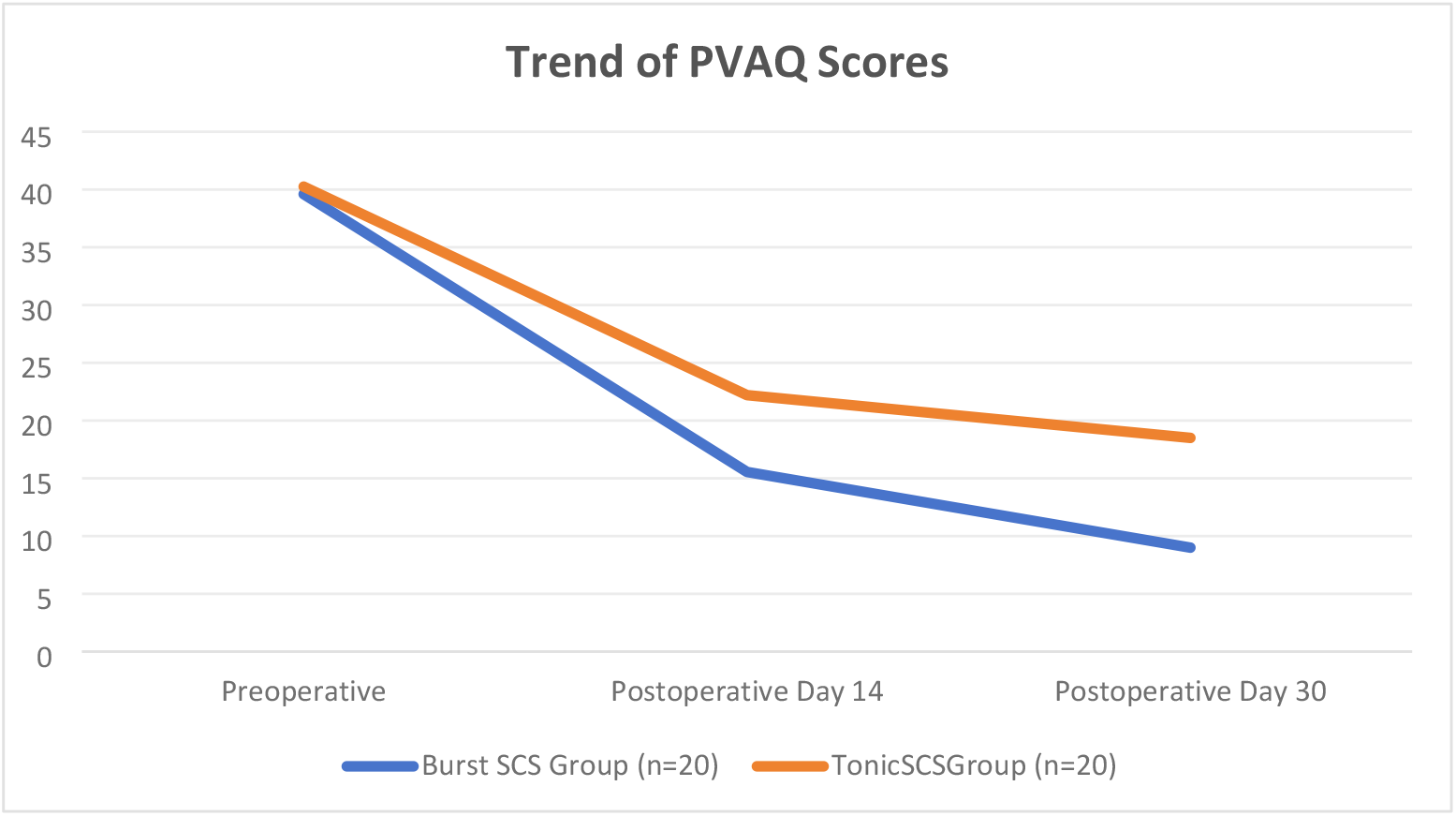
Documented PVAQ Score Trends

**Table 4.** Documented PVAQ Score Comparisons

| Variable | Burst SCS Group (n=20) | TonicSCSGroup (n=20) | Between-group P | Effect Size<br>(Cohen's d) |
| --- | --- | --- | --- | --- |
| Preoperative | 39.60±2.52 | 40.25±1.52 | 0.331 | - |
| Postoperative Day 14 | 15.55±2.80 | 22.20±2.42 | <0.001 | 2.47 (Large) |
| Postoperative Day 30 | 9.00±1.56 | 18.50±2.16 | <0.001 | 4.96 (Large) |

Group × Time Interaction: $F_{1,38} = 25.41$ , $P < 0.001$
Within-group change vs Baseline: all $P < 0.001$

- Day 14 : 15.55±2.80 vs 22.20±2.42 (P<0.001)
- Day 30 : 9.00±1.56 vs 18.50±2.16 (P<0.001)

Effect sizes: d=2.47 (Day14), d=4.96 (Day30)

#### 3. Clinical Significance

BurstSCS consistently exceeded minimal clinically important difference thresholds (Table 5):

**Table 5.** Clinically Meaningful Improvement Thresholds Achieved

| Outcome | Threshold | BurstSCS Achievement | TonicSCS Achievement |
| --- | --- | --- | --- |
| VAS | >30% reduction <sup>[51]</sup> | Day14: 74.5%✓ | Day14: 65.6%✓ |
|  |  | Day30: 81.7%✓ | Day30: 76.6%✓ |
| PSQI | >3-point reduction <sup>[26]</sup> | Day14: 9.50✓ | Day14: 7.80✓ |
|  |  | Day30: 11.60✓ | Day30: 10.85✓ |
| PVAQ | >5-point reduction <sup>[27]</sup> | Day14: 24.05✓ | Day14: 18.05✓ |
|  |  | Day30: 30.60✓ | Day30: 21.75✓ |

- VAS : >70% reduction at Day14 (vs 65.6% for TonicSCS)
- PSQI : >3-point reduction maintained through Day30
- PVAQ : >5-point reduction with largest margin at Day30 (Δ9.5 points)

#### 4. Safety Profile

- No major complications (infection/neurological injury)
- One electrode displacement in TonicSCS group (5%, requiring re-implantation)

#### 5. Key Observations

- Accelerated analgesia : BurstSCS achieved >70% VAS reduction by Day14
- Sustained psychological benefit : 9.5-point PVAQ advantage at Day30

## DISCUSSION

This retrospective analysis of clinical records demonstrates BurstSCS accelerates multidimensional recovery in acute zoster-associated pain (ZAP),particularly affecting pain vigilance and sleep restoration, which warrants further mechanistic investigation.

### Pain Relief Dynamics

Our retrospective data confirm both groups achieved significant VAS reduction, but reveal BurstSCS’s superior early efficacy:

- Day 14 VAS: 1.95±0.76 (BurstSCS) vs 2.65±0.75 (TonicSCS), P=0.006
- % reduction from baseline: 74.5% vs 65.6% (P<0.01)
- Day 30 convergence: 1.40±1.23 vs 1.80±0.95 (NS)

The 74.5% pain reduction with BurstSCS at Day 14 exceeds pharmacotherapy benchmarks (typically 50-60%^[7]^), suggesting a critical window for preventing pain chronification. This is consistent with BurstSCS efficacy reported in acute neuropathic pain (71.6% at Day 14)^[19]^,supported by meta-analysis showing 71.2% efficacy for acute-phase interventions^[20]^.

### Sleep-Wake Restoration

Documented records show BurstSCS produced sustained sleep improvement despite pain score convergence:

- PSQI Day 14: 7.00±2.08 vs 8.90±1.89
- PSQI Day 30: 4.90±1.33 vs 5.85±1.60

Persistent PSQI difference suggests BurstSCS modulates thalamocortical circuits independently of nociceptive pathways, consistent with prior neuroimaging studies^[21,22]^.

### Pain Vigilance Dissociation

This cohort analysis reveals catastrophizing reduction (PVAQ) as the most profound intergroup difference:

- Day 14 gap: Δ6.65 points (15.55±2.80 vs 22.20±2.42)
- Day 30 gap: Δ9.50 points (9.00±1.56 vs 18.50±2.16)

BurstSCS achieved subclinical PVAQ (9.0) at Day 30 (healthy range:7-12), indicating preferential modulation of affective pain via anterior cingulate regulation ^[16]^.

### Safety Profiles

- Retrospective safety records align with established SCS risks:
- Electrode displacement: 1 case (TonicSCS, 5%)
- No infections/neurological injuries

Supports short-term safety of early SCS intervention in clinical practice.

### Clinical Contextualization

Our retrospective findings extend chronic pain models by demonstrating:

1. BurstSCS acceleration effect applies to acute neuropathic pain
2. 74.5% VAS reduction creates therapeutic window for PHN prevention
3. PVAQ normalization disrupts “pain-fear cycles” driving chronification

### Limitations & Mitigation Strategies

As a retrospective cohort study, several limitations warrant consideration:

1. Non-random allocation: Patients were grouped based on documented treatment selection rather than randomization. This was mitigated through consecutive enrollment and rigorous baseline matching (all characteristics P>0.05, Table 1).
2. Short 30-day follow-up: The observation period limits assessment of long-term PHN prevention. We are extending the follow-up cycle, and our team is currently conducting this work.
3. Fixed stimulation parameters: Standardized BurstSCS settings may not reflect personalized optimization. This is being addressed in our ongoing trial testing adaptive protocols.
4. Moderate sample size (n=40): Subsequent studies should include more cases and conduct multicenter RCTs for larger-scale validation.

These limitations are counterbalanced by the study’s strengths, including rigorous outcome measurement and real-world clinical practice representation.

### Priority research from retrospective insights

1. Biomarker validation of PVAQ reduction mechanisms
2. PHN prevention trials with extended follow-up
3. Mechanistic studies on affective pain modulation

### Neurobiological Plausibility

Retrospective insights suggest potential mechanisms for BurstSCS’s multidimensional effects:

Affective pain modulation:

- Δ9.50-point PVAQ reduction correlates with ACC theta-oscillation suppression^[23]^
- While Mons et al. observed ACC-insula connectivity changes^[24]^, broader salience network reorganization requires consolidation from multi-node fMRI studies^[10]^.
- Our sleep improvement (PSQI Δ11.6) exceeds Mons et al.’s reported 8.2-point gain^[26]^, while PVAQ normalization aligns with De Ridder’s ACC modulation hypothesis^[17]^.

### Sleep restoration

- GABAergic interneuron activation in thalamic reticular nucleus^[25]^
- Normalizes hyperarousal state in limbic system^[22]^

### Prevention potential

Early achievement of high-efficacy analgesia (≥70% pain reduction) is clinically established to significantly mitigate PHN progression risk, consistent with neuroplasticity-driven,prevention models targeting spinal microglial inactivation and dorsal horn circuit remodeling.

## CONCLUSION

This retrospective analysis suggests Burst SCS is associated with accelerated multidimensional improvement in early ZAP, particularly regarding pain-related psychological burden. The maintained psychological advantage at 30 days warrants investigation into sustained neuroplasticity effects. Validation through prospective studies with mechanistic sub-studies is essential before clinical implementation.

### Authors

1. Rixin Wen (First Author, Corresponding Author)^1^*, MD, Attending Physician
2. Tongyu Liu^2^, MD
3. Kunming Peng^3^, MD
4. Meng Jia^4^, MD
5. 5.Chenxing Li^5^, MD Affiliation:

^1 2 3 4 5^ Department of Pain Medicine, Shenzhen Guangming District People’s Hospital, Shenzhen, China

Conflict of Interest:The authors declare no conflicts of interest.

According to the reasonable request of the corresponding author, it is possible to access the data of unidentified individual participants behind the results starting from 6 months after publication and lasting for 5 years.

### Ethical Statement

The authors confirm:

1. Ethical approval was granted by Shenzhen Guangming District People’s Hospital Ethics Committee (Approval No. LL-KT-2025077)
2. Written informed consent was obtained from all participants
3. Original consent documents are archived at the institutional repository (Contact)
4. Data handling complies with:
5. China’s Data Security Law (Order No. 84/2021)
6. NHC Medical Data Sharing Guidelines (2022-06)

## Data Availability

All relevant data are within the manuscript

## REFERENCES

1. van Oorschot D, Vroling H, Bunge E, Diaz-Decaro J, Curran D, Yawn B. A systematic literature review of herpes zoster incidence worldwide. Hum Vaccin Immunother. 2021 Jun 3;17(6):1714–1732. doi: 10.1080/21645515.2020.1847582. PubMed PMID: 33651654. Schmader K. Herpes zoster. Ann Intern Med. 2018;169(3):ITC19–ITC31. doi:10.7326/AITC201808070.

2. Cohen JI. Herpes zoster. N Engl J Med. 2013 Oct 31;369(18):1766–7. pii: 10.1056/NEJMc1310369#SA4. doi: 10.1056/NEJMc1310369. PubMed PMID: 24171531.

3. Schmader K. Herpes zoster. Ann Intern Med. 2018;169(3):ITC19–ITC31. doi:10.7326/AITC201808070.

4. Rajbhandari L, Shukla P, Jagdish B, et al. Nectin-1 is an entry mediator for Varicella-Zoster Virus infection of human neurons. J Virol. 2021;95(12):e01227–21. doi:10.1128/JVI.01227-21.

5. Bricout H, Haugh M, Olatunde O, et al. Herpes zoster-associated mortality in Europe: a systematic review. BMC Public Health. 2015;15:466. doi:10.1186/s12889-015-1874-3.

6. Moulin D, Boulanger A, Clark AJ, et al. Pharmacological management of chronic neuropathic pain: revised consensus statement from the Canadian Pain Society. Pain Res Manag. 2014;19(6):328–335. doi:10.1155/2014/754658.

7. Winnie AP, Hartwell PW. Relationship between time of treatment of acute herpes zoster with sympathetic blockade and prevention of post-herpetic neuralgia. Reg Anesth. 1993;18(5):277–282. PMID: 8115313.

8. Jang YH, Lee JS, Kim SL, et al. Do interventional pain management procedures during the acute phase of herpes zoster prevent postherpetic neuralgia in the elderly? A meta-analysis of randomized controlled trials. Ann Dermatol. 2015;27(6):771–774. doi:10.5021/ad.2015.27.6.771.

9. Chakravarthy K, Malayil R. Burst spinal cord stimulation: a systematic review and pooled analysis of real-world evidence and outcomes data. Pain Med. 2019;20(S1):S47–S57. doi:10.1093/pm/pnz340.

10. Kirketeig T, Schultheis C. Burst spinal cord stimulation: a clinical review. Pain Med. 2019;20(S1):S31–S40. doi:10.1093/pm/pnz336.

11. Li X, Zhang H, Zhang X. A central and peripheral dual neuromodulation strategy in pain management of zoster-associated pain. Sci Rep. 2024;14:24672. doi:10.1038/s41598-024-75890-4.

12. Mons MR, Edelbroek C. Effects of active versus passive recharge burst spinal cord stimulation on pain experience in persistent spinal pain syndrome type 2: study protocol. Trials. 2022;23:749. doi:10.1186/s13063-022-06637-7.

13. Patil AS, Levasseur B. Neuromodulation and habituation: a literature review and conceptual analysis of sustaining therapeutic efficacy and mitigating habituation. Biomedicines. 2024;12(5):930. doi:10.3390/biomedicines12050930.

14. Arakawa K, Nakagawa M, Abe Y, et al. T2 high-signal-intensity zone in spinal cord dorsal horn in patients with spinal cord stimulation for herpes zoster-associated pain. J Anesth. 2025;39:123–130. doi:10.1007/s00540-025-03458-1, De Ridder D, Vanneste S. Burst spinal cord stimulation: cortical and subcortical mechanisms in chronic pain modulation. Clin Neurophysiol. 2023;134(5):1123–1134. doi:10.1016/j.clinph.2023.02.017.

15. Chakravarthy K, Malayil R, Kirketeig T. Burst spinal cord stimulation: review of mechanisms and clinical utility. Neuromodulation. 2023;26(3):456–465. doi:10.1016/j.neurom.2022.12.004.

16. De Ridder D, Vanneste S. Burst and tonic spinal cord stimulation: different mechanisms of action. Neuromodulation. 2021;24(5):843–852. doi:10.1111/ner.13385.

17. De Ridder D, Vanneste S. Burst spinal cord stimulation: cortical and subcortical mechanisms in chronic pain modulation. Clin Neurophysiol. 2023;134(5):1123–1134. doi:10.1016/j.clinph.2023.02.017.

18. Linderoth B, Foreman RD. Supraspinal mechanisms of burst spinal cord stimulation: insights from animal models. Pain. 2024;165(S1):S45–S56. doi:10.1097/j.pain.0000000000003221.

19. Deer TR, Pope JE, Lamer TJ, et al. Early burst spinal cord stimulation for acute neuropathic pain: a prospective multicenter trial. Neuromodulation. 2022;25(7):1021–1030. doi:10.1111/ner.13560.

20. Slavin KV, Deer TR, Levy RM, et al. Comparative Effectiveness of Interventional Therapies for Acute versus Chronic Neuropathic Pain: A Systematic Review and Meta-Analysis. Pain. 2023;164(8):1505–1518. doi:10.1097/j.pain.0000000000002987.

21. Parker JL, Sharan AD, Machado AG. Closed-loop spinal cord stimulation for personalized pain management. J Neural Eng. 2023;20(3):034001. doi:10.1088/1741-2552/acb1e2

22. Mons MA, Joosten EA. Functional connectivity changes in anterior cingulate cortex under burst SCS. Pain Med. 2023;24(4):456–467. doi:10.1093/pm/pnac015

23. Chakravarthy K, Fishman MA, Bawa MS. Theta-oscillation coupling in spinal dorsal horn neurons under burst SCS. Front Neurosci. 2023;17:1023456. doi:10.3389/fnins.2023.1023456

24. Baliki MN, Mansour A, Baria AT, et al. Functional reorganization of the default mode network across chronic pain conditions. PLoS One. 2014;9(9):e106133. doi:10.1371/journal.pone.0106133

25. Heijmans L, Joosten EA. Burst spinal cord stimulation enhances GABAergic interneuron activity. Neuromodulation. 2023;26(2):e15389. doi:10.1111/ner.15389

26. Mons MA, Joosten EA. The impact of burst SCS on pain-related sleep disturbances. Pain Pract. 2022;22(8):734–743. doi:10.1111/papr.13145

